# Longitudinal associations between adverse childhood experiences with moderate-risk to problem gambling in young adulthood: A prospective UK cohort study

**DOI:** 10.64898/2026.04.02.26349298

**Authors:** Emily Patterson, Raffaello Rossi, Hannah Sallis, Emma Dennie, Laura D Howe, Alan Emond, Annie Herbert

## Abstract

Previous research links Adverse Childhood Experiences (ACEs) with problem gambling, but most studies rely on retrospective reporting and focus narrowly on maltreatment, overlooking adversities such as parental mental health issues.

Using data on 3794 young adults in the Avon Longitudinal Study of Parents and Children, we examined longitudinal associations between 10 prospectively measured ACEs (individually and cumulatively), and moderate-risk/problem gambling (Problem Gambling Severity Index >=3) at ages 17, 20 and 24, adjusted for socioeconomic and other background factors. Population attributable fractions (PAFs) estimated proportions of cases potentially attributable to ACEs.

Most ACEs were associated with higher odds of moderate-risk/problem gambling across ages (24/30 estimates) after adjustment, though effect sizes were generally small (median adjusted odds ratio [aOR] 1.31, interquartile range 1.24-1.59), and confidence intervals (CIs) wide. Sexual abuse showed the strongest association (aORs 2.4-4.2, CIs 0.5-10.5), while bullying and parental conviction were associated at ages 17 and 20 only, parental separation age 24 only. Evidence for a dose-response relationship was weak. PAFs suggested ACEs accounted for up to 12% of moderate-risk/problem gambling cases.

These findings highlight potential impacts of ACEs on later gambling behaviour, but imprecise estimates suggest findings should be interpreted cautiously and strengthened through larger datasets and meta-analyses.

## 1. Introduction

Problem gambling^1^ (Blaszczynski et al., 2020), defined as gambling behaviour that disrupts personal, family, or social functioning, affects an estimated 2.5% of adults in Great Britain, with a further 3.7% at moderate risk (Gambling Commission, 2024). It is associated with a wide range of physical, mental, and social harms (Emond & Griffiths, 2020; Gunstone & Gosschalk, 2020; Public Health England & Office for Health Improvement and Disparities, 2023). These harms extend beyond the individual, with around 7% of the UK population affected by someone else’s gambling (Gunstone & Gosschalk, 2020; Public Health England & Office for Health Improvement and Disparities, 2023). Evidence also suggests an intergenerational dimension, highlighting and the importance of understanding early-life risk factors (Dowling et al.).

Young adults face may be particularly vulnerable to gambling harms due to developmental changes in impulsivity and decision-making, greater exposure to online gambling environments, and increased financial and social pressures (Blakemore & Choudhury, 2006; Chambers & Potenza, 2003; Delfabbro et al., 2006; Emond et al., 2020; NHS Digital, 2021; Public Health England & Office for Health Improvement and Disparities, 2023; Rossi & Nairn, 2024, 2025). Understanding early-life risk factors that contribute to gambling in this developmental window is therefore crucial.

One such early risk factor is Adverse Childhood Experiences (ACEs): stressful or traumatic events occurring before age 18, including maltreatment (being emotionally or physically abused or neglected, or sexually abused by an adult in the household), or household dysfunction (e.g. growing up with a caregiver who themselves are vulnerable through mental illness, substance misuse, or criminal activity) (Felitti et al., 1998). ACEs are common: around 50% of UK adults have reported experiencing at least one ACE and 13% report four or more (Ford et al., 2019), rising to 81% and 41% among those seeking treatment for gambling-related harm (Moxey et al., 2026). ACEs have been linked to a broad range of adverse health, social, and behavioural outcomes across the life-course (Bellis et al., 2019; Felitti et al., 1998), including addictive behaviours such as smoking and substance misuse. Proposed mechanisms include stress-related disruptions to emotional regulation and impulsivity (Bick & Nelson, 2016; Blakemore & Choudhury, 2006; Espeleta et al., 2018; Soares et al., 2021), which may plausibly increase vulnerability to gambling.

Despite growing interest, robust longitudinal evidence on ACEs and gambling is limited. Prior reviews have highlighted associations between child maltreatment and later problem gambling, but most available studies rely on cross-sectional or retrospective designs (Lane et al., 2016). These designs raise concerns about recall bias and temporal ambiguity, particularly when ACEs are reported after gambling problems have developed (Baldwin et al., 2019). More recent work examining broader stressful life events and family level influences similarly suggests that early adversity may play a role in gambling vulnerability, but these studies still lack prospective childhood ACE measurement(Russell et al., 2022; Smith et al., 2025). Existing studies have also tended to focus narrowly on maltreatment rather than the full possible range of ACEs; examining ACEs side-by-side could help clarify whether particular domains of adversity, or cumulative burden, are most relevant for later gambling behaviour.

Policy discussions in the UK have similarly highlighted gaps in the evidence: the 2023 Gambling Act White Paper concluded that there was insufficient evidence for a strong association between child maltreatment and gambling harms (Public Health England & Office for Health Improvement and Disparities, 2023). Stronger prospective evidence on ACE-gambling could support prevention strategies, early identification, and trauma-informed care (Asmussen et al., 2020), particular as gambling-related care increasingly moves into the National Health Service’s remit (National Institute for Health and Care Excellence, 2025).

## 2. Aims and objectives

Using data from the Avon Longitudinal Study of Parents and Children (ALSPAC) birth cohort we aimed to investigate the longitudinal associations between 10 different ACEs (as total number of ACEs and each ACE separately) measured during childhood, and moderate-risk or problem gambling in young adulthood at ages 17, 20, and 24 years. Our focus was on associations adjusted for measured background confounders, that is, the goal was to the best of our ability, to estimate the causal contribution of ACEs to subsequent gambling risk.

The specific objectives were to investigate:

1. the longitudinal relationship between ACEs (individual and number of) and moderate-risk gambling (a Problem Gambling Severity Index’ [PGSI] score of 3+) as measured by the PGSI.
2. the extent to which ACEs accounted for cases of moderate-risk or problem gambling in young adults

Given findings between ACEs and other addictive behaviours we hypothesised that the adjusted associations between the number of ACEs and experiencing moderate-risk or problem gambling would follow a ‘dose-response’, where there were stronger associations for higher numbers of ACEs (Hughes et al., 2017).

## 3. Methods

### 3.1 The ALSPAC study

Our study used data from the Avon Longitudinal Study of Parents and Children (ALSPAC), a UK-based birth cohort (www.bristol.ac.uk/alspac) (Boyd, Fraser, et al., 2013; Fraser et al., 2013; Northstone et al., 2019). Pregnant people with an expected delivery date between 1^st^ April 1991-31^st^ December 1992, and who lived in Avon, UK were invited to join the cohort. 20,248 pregnancies were identified as being eligible and the initial number of pregnancies enrolled was 14,541. Of the initial pregnancies, there was a total of 14,676 foetuses, resulting in 14,062 live births and 13,988 children who were alive at one year of age. When the oldest children were approximately seven years of age, an attempt was made to bolster the initial sample with eligible cases who had failed to join the study originally. The total sample size for analyses using any data collected after the age of seven is therefore 15,447 pregnancies, resulting in 15,658 foetuses. Of these 14,901 children were alive at one year of age, the index offspring or G1 cohort.

ALSPAC data were collected and managed using REDCap (Research Electronic Data Capture) hosted at the University of Bristol. REDCap is a secure, web-based software platform designed to support data capture for research studies (Harris et al., 2009). Please note that the study website contains details of all the data that is available through a fully searchable data dictionary and variable search tool (http://www.bristol.ac.uk/alspac/researchers/our-data/). At age 17 years, 5,217 ALSPAC participants completed a research clinic visit at which they were invited to fill in an additional questionnaire about gambling behaviours (including PGSI questions), to which 3,829 consented. At ages 20 and 24, 9,067 and 9215 cohort participants were still enrolled and contactable in ALSPAC; of these participants 2,625 and 2,758 completed online or postal questionnaires.

### 3.2 Current study participants

For this study, we analysed data on the children of the cohort (the index offspring or ‘G1s’). Participants were included in this study if they had data available on at least one ACE and from a completed PGSI at age 17, 20, and/or 24 years. Fig. 1 shows the numbers of participants that were originally in ALSPAC at the outset through to those included in the current study sample of 3,794. As part of rules for accessing ALSPAC data, triplets and quadruplets were excluded from analyses, as this was extremely rare (n=13) and these individuals are considered too identifiable (Boyd, Golding, et al., 2013). In the original ALSPAC cohort, there were 195 sets of twins; after applying all other exclusion criteria, there were 40 sets of twins (1% of the study sample) in the analytical sample – we posit that clustering within these families would have minimal impact on estimates.

**Fig. 1.**
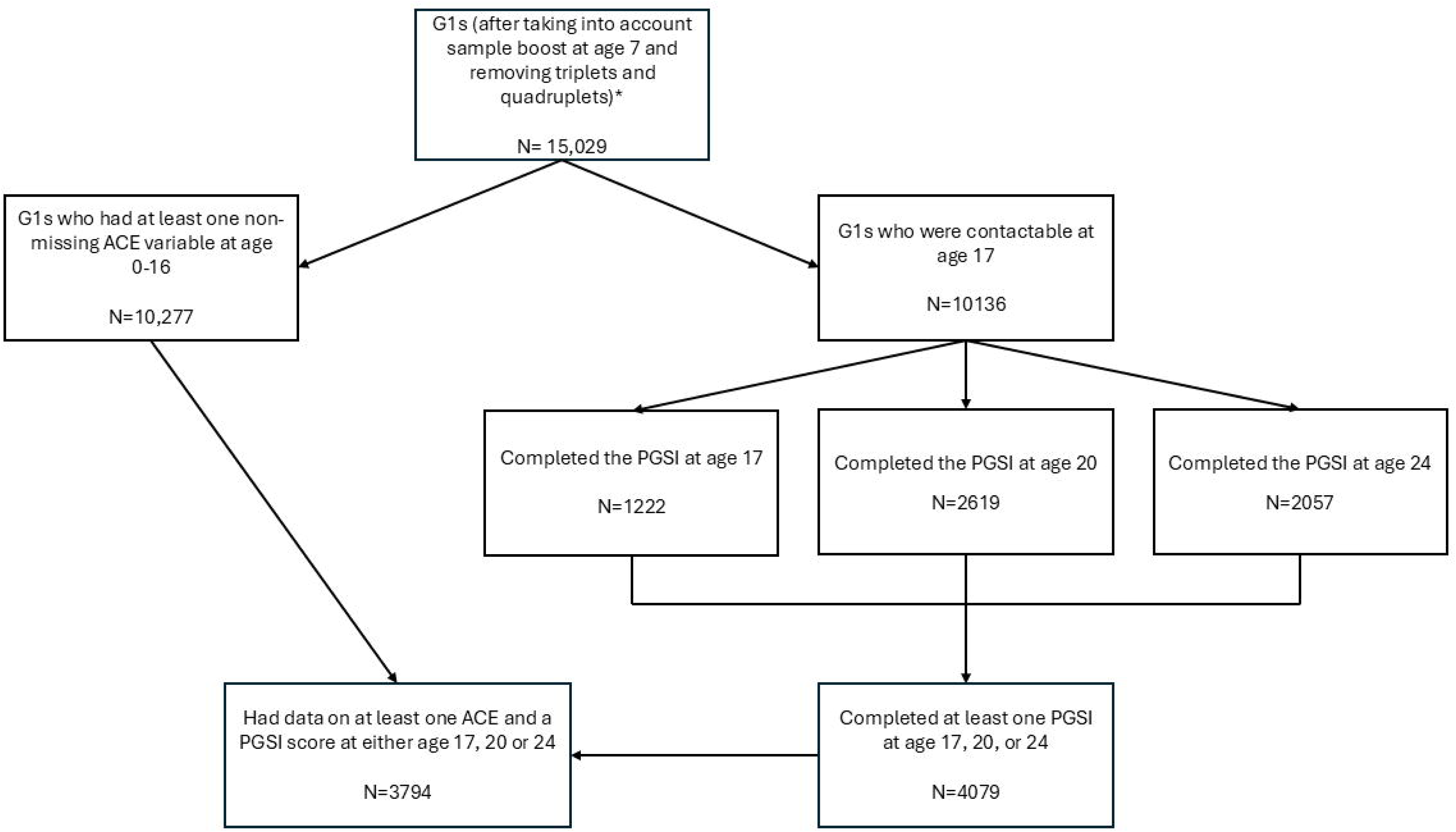
Flowchart of study participants. *Throughout, numbers are participants with non-missing sex

### 3.3 Exposure: Adverse Childhood Experiences

Individual binary variables (yes/no) for the following ACEs have been generated for the ALSPAC G1s previously: physical abuse, sexual abuse, emotional abuse, emotional neglect, bullying, violence between parents, household substance abuse, parental mental health problems or suicide, parent convicted of an offence, and parental separation. These ACEs are consistent with those listed in the World Health Organisation (WHO) Adverse Childhood Experiences International Questionnaire (ACE-IQ) (Lereya et al., 2015; World Health Organisation, 2020). These binary ACE variables have been derived using 541 questionnaire items from responses to survey and clinic questions that were parent-reported until the children were eight years old, both parent- and child-reported until age 16. Further details are provided in Houtepen *et al* (Houtepen et al., 2018). We updated these binary measures to include only prospective measures up to age 16 (to not include retrospective reporting of ACEs at ages 22-23). To both account for likely high levels of co-occurrence of ACEs for individuals, and to explore a potential dose-response relationship, a total ACE ‘score’ was generated by summing the total number of different ACEs experienced between 0 and 16 years (i.e. the score could range from 0 to 10). ACE counts were highly positively skewed; treating them as a continuous variable would imply linearity and produced unstable estimates at higher scores. We therefore categorised ACEs as 0, 1, 2-3, and 4+, following established practice (Hughes et al., 2017; Lacey & Minnis, 2019), and reflecting commonly used thresholds for low, moderate, and high cumulative adversity.

### 3.4 Outcome: Gambling risk categories

The primary outcome was moderate-risk or problem gambling, measured using the shortened nine-item PGSI questionnaire (Ferris J, 2001). The PGSI is a standardised measure of gambling problems, and each item (of nine) is measured on a four-point scale with a maximum score of 27. Typically, researchers sum across the nine items and categorise the PGSI score as the following: 0 = ‘No Problem Gambling’; 1-2 = ‘Low risk of Problem Gambling’; 3 - 7 = ‘Moderate-risk of Problem Gambling’; 8+ = ‘Problem Gambling’. with the number of participants meeting the threshold for problem gambling (scores of ≥8) was small at each age (n=7, 25, and 30 at age 17, 20, and 24, respectively. Therefore, we combined moderate-risk and problem gambling categories to create a binary outcome (a score of ≥3 vs. 0-2), consistent with other UK population surveys where high-severity cases are uncommon (Glasgow, 2024; NHS Digital, 2023), as it improves model stability while retaining the distinction between no/low-risk and at-risk gambling. The PGSI has demonstrated good internal consistency and reliability in UK population samples (e.g., Cronbach’s alpha = 0.89) (Glasgow, 2024), supporting its use as a categorical measure in epidemiological studies. To capture earlier stages of gambling risk that may precede moderate-risk gambling in young adulthood, we additionally examined an outcome defined as PGSI ≥ 1..

### 3.5 Covariates

We adjusted for the covariates that we hypothesised to be potential confounders of the ACE-gambling relationship. These covariates were all recorded at the time of the mother’s pregnancy or birth of the baby, and many representing family socio-economic background, a key potential confounder of the ACE-gambling association: biological sex (registered at birth; gender identity was not available from the data), parity (0, 1, 2+), mother’s and father’s highest educational qualification (A-level or higher vs. lower), mother’s social class (NS-SEC category, III manual+ or not, and mother ownership/mortgage (yes/no).

### 3.6 Missing data approach

We accounted for missing data using multiple imputation with chained equations. We fitted imputation models that included ACEs, PGSI categories (no; low; moderate; problem) at all time-points, all covariates included in the adjusted model and auxiliary variables (at time of mother’s pregnancy/young person’s birth: numeric variables for mother’s age, child’s birthweight, mother’s Edinburgh Postnatal Depression Score (EPDS) and partner’s EPDS; and binary variables for White ethnicity and mother smoking in pregnancy). We assumed data to be Missing At Random, which we feel to be plausible given large amounts of information from available data including auxiliary covariates. We fitted models separately for women and men, given likely different patterns of missingness by sex, and potential interactions between ACEs and sex, created twenty imputed datasets, and pooled estimates across imputed datasets using Rubin’s rules (Rubin, 1987). We present estimates after multiple imputation within the main results, and estimates from the complete data set only within supplementary materials.

### 3.7 Statistical Analysis

All data cleaning and statistical models were carried out in STATA v.17. Tables and figures were created using R v.4.3.1. We analysed PGSI outcomes at each of the three ages of 17, 20 and 24, separately, representing distinct stages of young adulthood. We considered three too few time-points to model within-person gambling trajectories or repeated-measures structures without imposing strong and unrealistic assumptions about developmental change.

Although our focus is on adjusted associations between ACEs and gambling, we also present crude associations within supplementary materials, for reference. We fitted binary logistic regressions to estimate the associations of individual ACEs and ACE scores, with moderate-risk+ gambling (vs. no-to-low-risk), as crude models, and adjusted for variables listed above under ‘*Covariates*’. We fitted 66 separate models [11 different ACE exposures (10 individual binary, one ACE score categorical), a binary outcome at three different time-points, each crude and adjusted]. We then repeated these models with the outcome of low-risk+ gambling, as a secondary outcome of interest.

Conclusions on the strength of evidence for associations were based on the point estimates of the estimated odds ratios (ORs) and their associated 95% confidence intervals (CIs). That is, we do not focus on ‘statistical significance’, as recommended by the statistical community.(Amrhein et al., 2019)

To explore the possibility of different relationships between ACEs and gambling between men and women, we included an interaction term between sex and the exposures (ACE score and individual ACEs). Most of these interaction terms had high p-values (median across 36 p-values =0.46, inter-quartile range=0.28 to 0.76). Therefore, in our final analyses and reporting we do not stratify models by sex.

To estimate the extent to which ACEs accounted for moderate-risk or problem gambling (anyone with a score of 3+ on the PGSI) in young adults, population attributable fractions (PAFs) were calculated based on relative risks approximated to estimated adjusted odds ratios (aORs). The PAFs can be interpreted as the maximum proportion of cases of at least moderate-risk gambling that could be attributed to experience of the specific ACE. PAFs were estimated using the formula described in **Supplementary Box S1**.

### 3.8 Ethics statement

Ethical approval for the study was obtained from the ALSPAC Ethics and Law Committee and the Local Research Ethics Committees (further details here: https://www.bristol.ac.uk/media-library/sites/alspac/documents/governance/Research_Ethics_Committee_approval_references.pdf). Informed consent for the use of data collected via questionnaires and clinics was obtained from participants following the recommendations of the ALSPAC Ethics and Law Committee at the time.

## 4. Results

### 4.1 Participants

A total of 3,794 participants had data on at least one ACE and had completed the PGSI at either ages 17, 20, or 24 years (2,079 female, 1,715 male) (**Table 1**). Most participants were White. Around 82% of participants’ maternal homes were owned or purchased with a mortgage, and around half of the participants’ parents had A-level qualifications. The median EPDS score in participants’ mothers was 6, with half having a score of 3 to 9 (representing from no to mild depression, to mild depression). Compared with the originally recruited ALSPAC cohort, participants were more likely to be White (95% vs. 93%), female (55% vs. 49%), and socioeconomically more advantaged (e.g. home ownership 82% vs. 69%).

**Table 1.** Breakdown of characteristics for participants with observed ACE and PGSI score, n (%) or median (inter-quartile range)^a^.

| Ethnic group | Study sample |  | Original ALSPAC cohort |  |
| --- | --- | --- | --- | --- |
| White | 3597 | (94.8) | 13973 | (93) |
| Other | 197 | (5.2) | 1056 | (7.0) |
| Sex |  |  |  |  |
| Female | 1715 | (45.2) | 7683 | (51.1) |
| Male | 2079 | (54.8) | 7346 | (48.9) |
| Maternal home ownership |  |  |  |  |
| Mortgaged/owned | 667 | (17.8) | 4609 | (31.1) |
| Rented or housing association | 3080 | (82.2) | 10219 | (68.9) |
| Social class (maternal) |  |  |  |  |
| I-III | 538 | (16.2) | 2621 | (21.4) |
| IV-V | 2783 | (83.9) | 9611 | (78.5) |
| Parity |  |  |  |  |
| 0 | 1753 | (48.5) | 6430 | (45.8) |
| 1 | 1268 | (35.1) | 4832 | (34.4) |
| 2+ | 604 | (16.7) | 2785 | (19.8) |
| Maternal smoking in pregnancy |  |  |  |  |
| No | 3180 | (84.0) | 11480 | (76.7) |
| Yes (1 or more cigarettes) | 606 | (16.0) | 3490 | (23.3) |
| Mother's highest educational qualification |  |  |  |  |
| <A-level | 2090 | (55.1) | 10005 | (66.6) |
| A-level+ | 1704 | (44.9) | 5024 | (33.4) |
| Father's highest educational qualification |  |  |  |  |
| <A-level | 1473 | (41.6) | 6763 | (51.1) |
| A-level+ | 2062 | (58.3) | 6438 | (48.6) |
| Mother age at delivery | 29 | (26 to 32) | 28 | (25 to 31) |
| Mother Edinburgh postnatal depression score | 6 | (3 to 9) | 7 | (3 to 10) |
| Father Edinburgh postnatal depression score | 3 | (1 to 6) | 4 | (2 to 7) |
| Adverse Childhood Experience |  |  |  |  |
| (age 0-16) |  |  |  |  |
| ACE score |  |  |  |  |
| 0 | 780 | (20.6) | 2680 | (17.8) |
| 1 | 1022 | (26.9) | 3853 | (25.6) |
| 2-3 | 1350 | (35.6) | 5539 | (36.9) |
| 4+ | 642 | (16.9) | 2957 | (19.7) |
| ACE experienced |  |  |  |  |
| Physical abuse | 702 | (18.5) | 2261 | (15.0) |
| Sexual abuse | 175 | (4.6) | 497 | (3.3) |
| Emotional abuse | 707 | (18.6) | 2840 | (18.9) |
| Emotional neglect | 694 | (18.3) | 3152 | (21.0) |
| Bullying | 913 | (24.1) | 3691 | (24.6) |
| Violence between parents | 749 | (19.7) | 3327 | (22.1) |
| Household substance abuse | 424 | (11.2) | 2186 | (14.5) |
| Parental mental health problems<br>or suicide | 1612 | (42.5) | 7042 | (46.9) |
| Parent convicted of an offence | 288 | (7.6) | 1384 | (9.2) |
| Parental separation | 1039 | (27.4) | 5128 | (34.1) |
| PGSI gambling categories at age<br>17 |  |  |  |  |
| Non-problem gambler (score 0) | 2802 | (73.8) | 10660 | (70.9) |
| Low-risk gambler (score 1-2) | 757 | (20.0) | 3283 | (21.8) |
| Moderate-risk gambler (score 3-7) | 208 | (5.5) | 926 | (6.2) |
| 8+ Problem gambler (score >=8) | 27 | (0.7) | 159 | (1.1) |
| PGSI gambling categories at age<br>20 |  |  |  |  |
| Non-problem gambler (score 0) | 2691 | (70.9) | 10102 | (67.2) |
| Low-risk gambler (score 1-2) | 836 | (22) | 3621 | (24.1) |
| Moderate-risk gambler (score 3-7) | 226 | (6.0) | 1086 | (7.2) |
| 8+ Problem gambler (score >=8) | 41 | (1.1) | 220 | (1.5) |
| PGSI gambling categories at age<br>24 |  |  |  |  |
| Non-problem gambler (score 0) | 2932 | (77.3) | 11130 | (74.1) |
| Low-risk gambler (score 1-2) | 611 | (16.1) | 2648 | (17.6) |
| Moderate-risk gambler (score 3-7) | 184 | (4.8) | 913 | (6.1) |
| 8+ Problem gambler (score $\geq 8$) | 67 | (1.8) | 338 | (2.2) |

- a. These descriptive statistics have been derived in each of the imputed datasets and averaged.

The prevalence of ACEs was high, with over three-quarters of ALSPAC participants having experienced at least one ACE (**Table 1**). The most frequently reported ACE was parental mental health problems or suicide, affecting 43% of participants, whereas the least reported ACE was sexual abuse, experienced by 5% of participants.

PGSI categories varied across ages. Across all three ages (17, 20, and 24 years), 71-77% were categorised as no-risk gamblers, 16-22% were low-risk, 5-6% were moderate-risk, and 1-2% were categorised as problem gamblers.

### 4.2 Association between ACEs and moderate-risk or problem gambling

A majority of ACEs had point estimates in the direction of increased odds of moderate-risk or problem gambling at ages 17, 20, and 24, with those exposed having up to 2.03 times the odds of moderate-risk+ gambling as those unexposed, but most effects generally small (28/30 estimates were positive ORs, median of 24 estimates 1.27, interquartile range [IQR] 1.10 to 1.36) (**Fig. 2**). See **Supplementary Table S1** for crude (unadjusted) ORs and CIs. These associations should be interpreted cautiously given wide confidence intervals, many of which included the null.

**Fig. 2.**
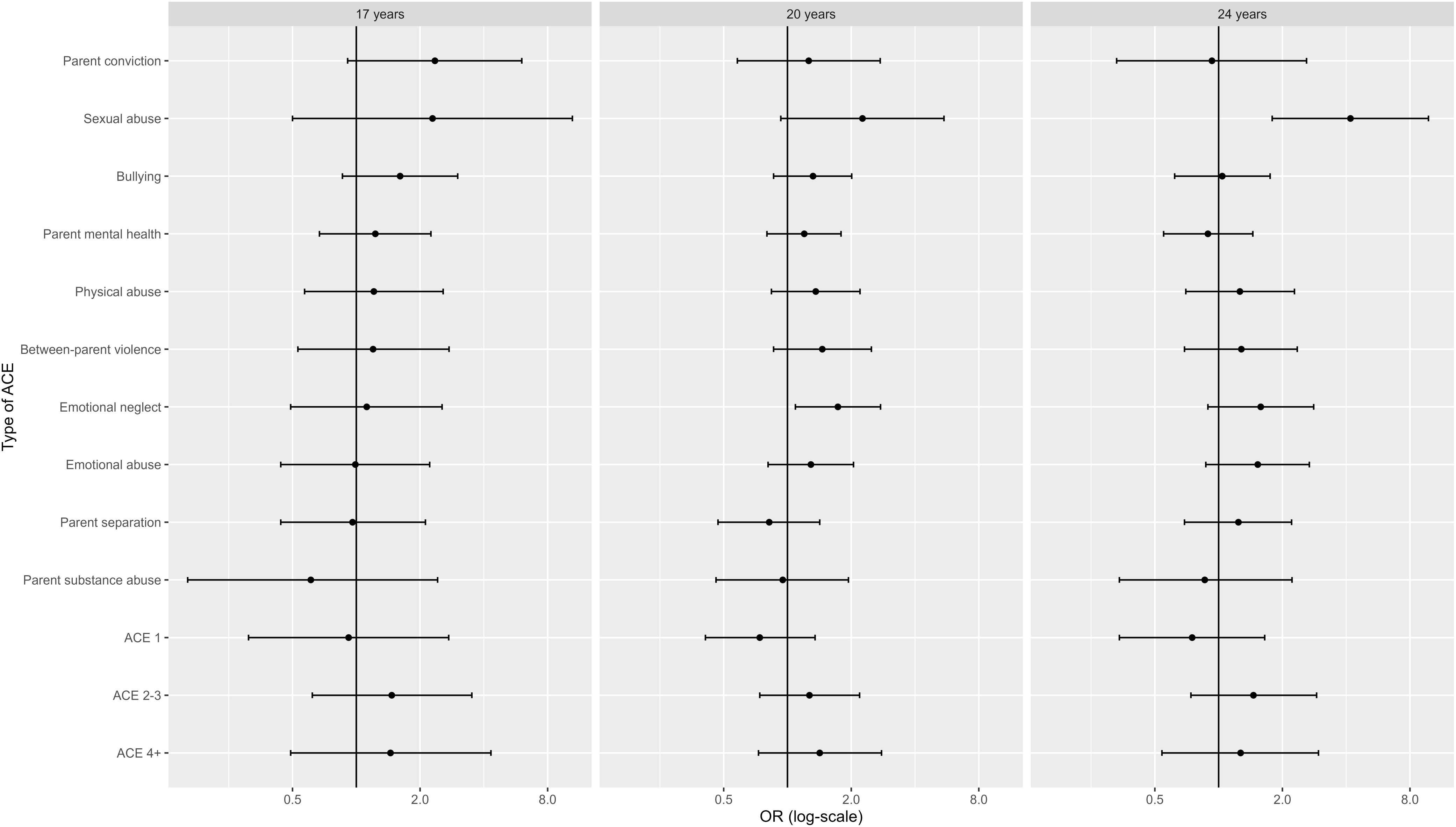
Adjusted odds ratios and confidence intervals for the relationship between Adverse Childhood Experiences and moderate-risk gambling at ages 17, 20, and 24^a^. a. Exact values underlying this figure are provided in **Supplementary Table S2**

After adjusting estimates for background socioeconomic factors, experiencing sexual abuse, physical abuse, between-parent violence, or emotional neglect were indicatively associated with higher odds of living with moderate-risk+ gambling at all age-points, (aORs ranged from 1.12 to 4.19; **Fig. 3, Table S2**), but CIs were wide, ranging from 0.49 to 10.47 for these ACEs, meaning that the true effect could plausibly be small or even absent. Of 30 possible estimates (10 ACEs, 3 outcome time-points), 24 were positive; again effect sizes were generally small (median aOR of these 24 estimates: 1.31, IQR 1.24 to 1.59). Sexual abuse yielded the largest point estimates, although the wide confidence intervals mean these findings remain highly uncertain (aOR at age 17: 2.29 [0.50 to 10.47]; age 20: 2.26 [0.93 to 5.48]; age 24: 4.19 [1.79 to 9.78]).

**Fig. 3.**
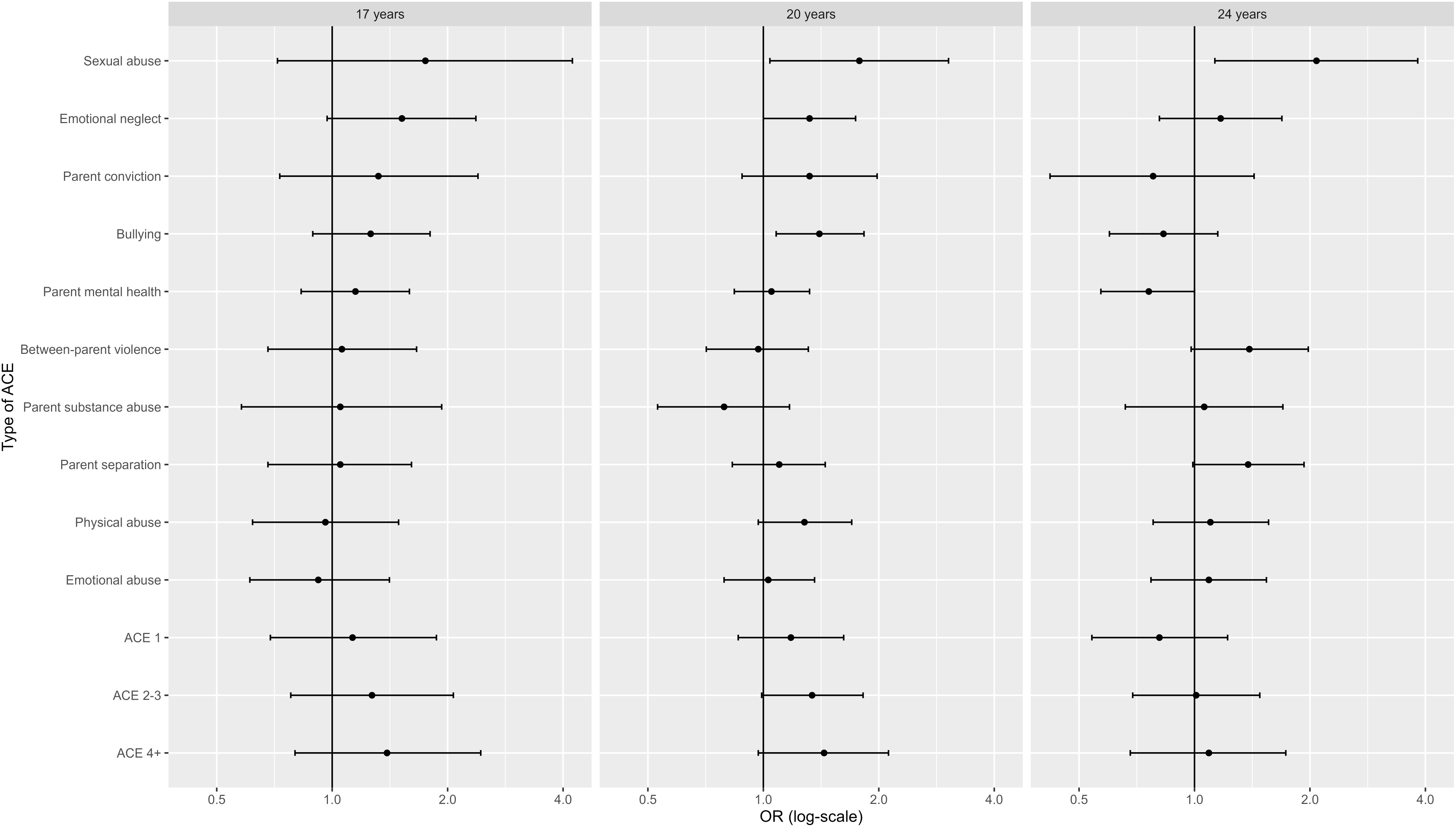
Adjusted odds ratios and confidence intervals for the relationship between Adverse Childhood Experiences and at-risk gambling at ages 17, 20, and 24^a^. a. Exact values underlying this figure are provided in **Supplementary Table S4**

In the case of parental conviction, bullying, and parental mental health problems, point estimates indicated higher odds of moderate-risk or problem gambling at ages 17 and 20 (aORs ranged 1.20 to 2.35), but not at age 24 (0.89 to 1.04), but again noting wide confidence intervals (0.33 to 6.04; **Fig. 3, Table S2**). There was little evidence for an association for emotional abuse, parental separation, or parental substance abuse, particularly at earlier ages (these aORs at age 17 ranged 0.61 to 0.99, their CIs ranged 0.16 to 2.42).

For number of ACEs, there were positive associations between experiencing 2-3 or 4+ ACEs vs. no ACEs, but with point estimates being similar between 2-3 and 4+ ACEs (aORs ranged 1.27 to 1.47), and CIs spanning the null (0.49 to 4.32). That is, there was little evidence for a dose-response relationship (**Fig. 3, Table S2**).

### 4.3 Population attributable fractions

The adjusted PAFs were low for all ACEs, with wide CIs (**Table 3**). The highest PAFs were for bullying and emotional neglect, where bullying accounted for 1%, 7%, and 12% of cases of moderate-risk or problem gambling at ages 17, 20, and 24 (CIs ranging −3% to 20%) and emotional neglect account for 2% to 9% (0% to 13%), reflecting that PAFs represent not just the size of the aOR, but also the prevalence of the ACE in question (bullying and emotional neglect was considered present for 24% and 18% of the study sample; **Table 1**).

**Table 3.**
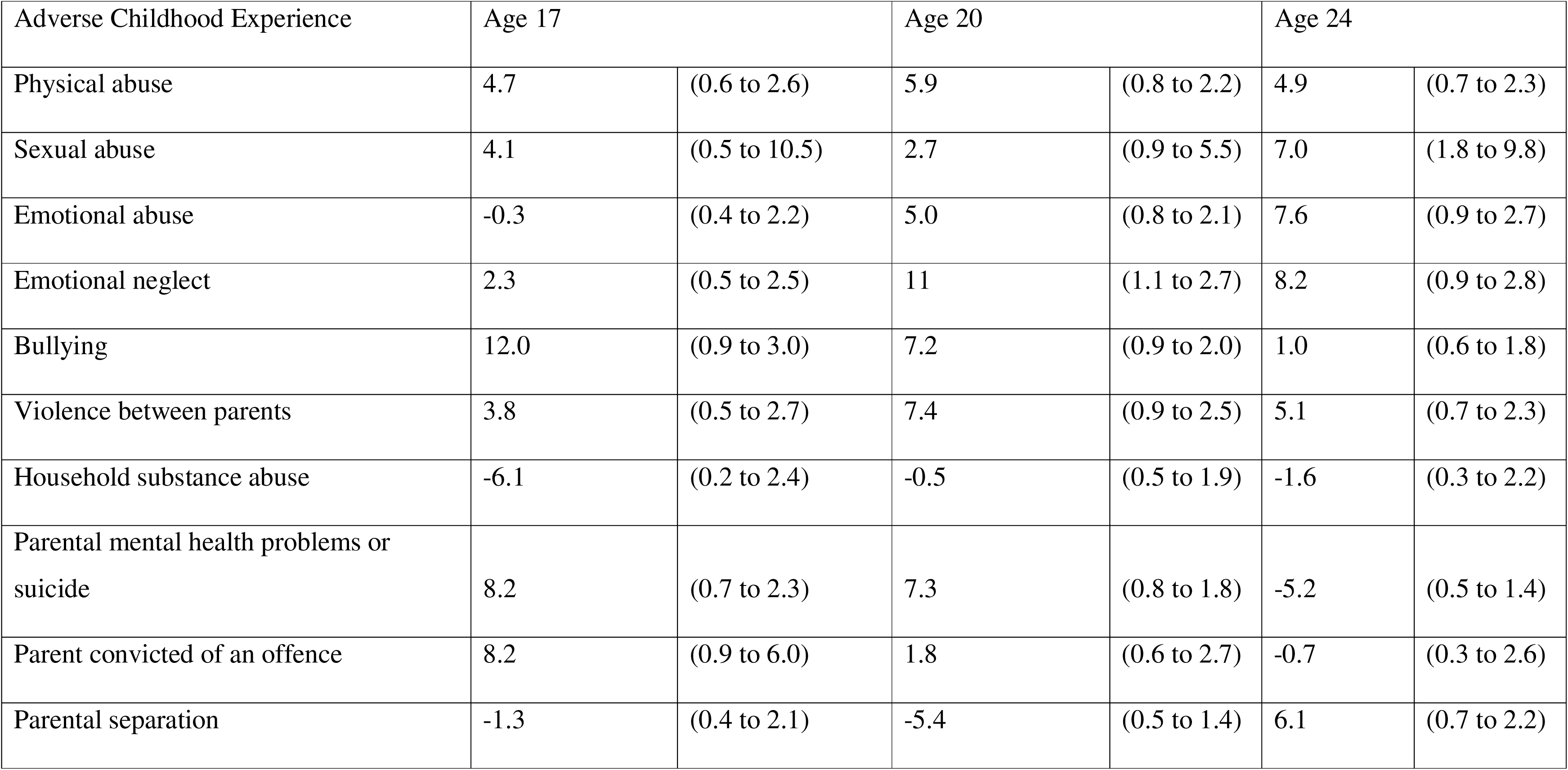
Population Attributable Fractions (PAFs) for individual Adverse Childhood Experiences and moderate-risk gambling at age 17, 20 and 24 years.

| Adverse Childhood Experience | Age 17 |  | Age 20 |  | Age 24 |  |
| --- | --- | --- | --- | --- | --- | --- |
| Physical abuse | 4.7 | (0.6 to 2.6) | 5.9 | (0.8 to 2.2) | 4.9 | (0.7 to 2.3) |
| Sexual abuse | 4.1 | (0.5 to 10.5) | 2.7 | (0.9 to 5.5) | 7.0 | (1.8 to 9.8) |
| Emotional abuse | -0.3 | (0.4 to 2.2) | 5.0 | (0.8 to 2.1) | 7.6 | (0.9 to 2.7) |
| Emotional neglect | 2.3 | (0.5 to 2.5) | 11 | (1.1 to 2.7) | 8.2 | (0.9 to 2.8) |
| Bullying | 12.0 | (0.9 to 3.0) | 7.2 | (0.9 to 2.0) | 1.0 | (0.6 to 1.8) |
| Violence between parents | 3.8 | (0.5 to 2.7) | 7.4 | (0.9 to 2.5) | 5.1 | (0.7 to 2.3) |
| Household substance abuse | -6.1 | (0.2 to 2.4) | -0.5 | (0.5 to 1.9) | -1.6 | (0.3 to 2.2) |
| Parental mental health problems or suicide | 8.2 | (0.7 to 2.3) | 7.3 | (0.8 to 1.8) | -5.2 | (0.5 to 1.4) |
| Parent convicted of an offence | 8.2 | (0.9 to 6.0) | 1.8 | (0.6 to 2.7) | -0.7 | (0.3 to 2.6) |
| Parental separation | -1.3 | (0.4 to 2.1) | -5.4 | (0.5 to 1.4) | 6.1 | (0.7 to 2.2) |

### 4.4 Findings for low-risk+ gambling compared to moderate-risk+

Findings for individual ACEs were similar when the threshold for the outcome was lowered to being at least low-risk of problem gambling (PGSI score of 1+ rather than 3+). Experiencing sexual abuse or emotional neglect were still associated with higher odds of living with at least low-risk gambling at all age-points after adjustment, and with generally similar magnitude (aORs ranged 1.23 to 2.09) (**Fig. 3, Tables S3 and S4)**. Parental conviction, bullying, and parental mental health problems, were also, as before, positively associated with low-risk+ gambling at ages 17 and 20 but not 24. The positive associations observed for between-parent violence or physical abuse and moderate-risk or problem gambling were not evident for at least low-risk gambling. There was stronger evidence for a dose-response relationship between number of ACEs and at least low-risk gambling than with moderate-risk or problem gambling (e.g. at age 17, aORs [CIs] increased from 0.92 [0.31 to 2.73] to 1.47 [0.62 to 3.51] to 1.45 [0.49 to 4.32] with increasing number of ACEs category), but this evidence was still weak given wide CIs. PAFs for low risk+ outcomes, which ranged up to 8% of low-risk+ cases at age 24 for emotional neglect, are presented in **Table S5**.

## 5. Discussion

Using prospective data from a large UK birth cohort, we observed suggestive positive but imprecise associations between several ACEs, particularly sexual abuse, physical abuse, and between-parent violence, and moderate-risk/problem gambling into young adulthood. There was little evidence for an increased number of different ACEs increasing the likelihood of moderate-risk or problem gambling. Population attributable fractions suggested that certain ACEs might account for up to 12% of cases of moderate-risk or problem gambling, but these estimates were highly uncertain due to wide confidence intervals.

### 5.1 Strengths and limitations

The main strengths of this study were prospective ACE measurement, reducing likelihood of recall bias, and repeated gambling outcomes. Consistency of associations across ages providing more robust evidence for a relationship between sexual abuse, physical abuse, between-parent violence and at least a moderate risk of problem gambling. However confidence intervals were wide and often spanned the null. Across almost all models, the width of the confidence intervals means that the precision of our estimates is limited, and the observed associations should therefore be interpreted as tentative rather than definitive. Multiple imputation minimised the impact of partially missing data and allowed us to include nearly 4,000 participants.

We captured ACEs through a comprehensive measure (captured using around 500 variables). Binary ACEs indicators were derived pragmatically from the available information on that construct and so there was variation between the individual ACEs in terms of what time periods were covered, and who the ACE was reported by (Houtepen et al., 2018). It is feasible that response accuracy is differential between those exposed to different ACEs due to the different thresholds for identification. For example, parental criminal conviction or separation are less likely to be susceptible to poor recall than emotional neglect. Further, ACEs such as emotional neglect at earlier time-points were parent-reported and thus may be under-reported due to social desirability bias. Such misclassification of ACEs would result in an under-estimate of positive associations; disproportionate misclassification between different types of ACEs would distort relative associations. We do not recommend ranking of ACEs in this study by magnitude of odds ratios, which slightly altered between moderate-risk+ and low-risk+ outcomes – nevertheless it is likely that sexual abuse is the most strongly associated with later gambling outcomes. Similarly, there is unlikely to be an association between emotional abuse and gambling outcomes at ages 17-24, given point estimates were close to the null at all three time-points for both risk thresholds. We found little evidence of a dose–response relationship between the number of ACEs and gambling risk, as point estimates for 2–3 and 4+ ACEs were similar and confidence intervals were wide. One possibility is that certain ACEs have stronger associations than others, meaning that a simple unweighted count may mask any true gradient. Alternatively, limited statistical power and measurement heterogeneity in ACE ascertainment may have attenuated any underlying dose–response pattern.

Although gambling was measured using a widely accepted and validated measure, the PGSI, there is still potential for reporting bias. Responses could be influenced by social desirability and therefore potential under-capture of gambling risk outcomes, which would again result in under-estimates of the positive association.

The generalisability of the findings may be limited by ALSPAC participant demographics, as the study sample were relatively affluent and majority White. Although the cohort originally enrolled in pregnancy was broadly representative of the British population at the time, differential attrition from ALSPAC has resulted in those who were socially disadvantaged being more likely to be lost to the study over time. Both ACEs and problem gambling are socially patterned, therefore we would hypothesise that any associations from this study are likely to be an underestimate (Felitti et al., 1998). However, the proportion of people experiencing ACEs and problem gambling within our study are within the range of previous prevalence estimates (Ford et al., 2019; Public Health England & Office for Health Improvement and Disparities, 2023). Further, previous methodological research in ALSPAC has found the effect of socially patterned loss-to-follow-up on risk estimates to be minimal (Howe et al., 2013).

### 5.2 Comparison with previous studies

Our point estimates for outcomes at multiple time-points were broadly in the same direction as previous cross-sectional studies linking sexual abuse to gambling harm (Allami et al., 2021; Bristow et al., 2022; Dennie et al., 2023; Poole et al., 2017), although the imprecision in our estimates means alignment should be interpreted cautiously. Other ACEs showed inconsistent associations: for example, an association was found between parental conviction, bullying, or parental mental health problems and moderate-risk of problem gambling at ages 17 and 20 but not 24, which correspond with previous findings of long-term effects of ACEs manifesting at different timepoints for other outcomes (Crick et al., 2022). However, given high levels of imprecision, and large number of models fitted as part of this study, increasing the likelihood of false positives. Replication in a large sample size than the current study would likely require cross-cohort studies and meta-analyses.

### 5.3 Implications for policy, practice, and research

This study provides tentative longitudinal evidence that supports, but does not confirm, a possible relationship between sexual abuse and problem gambling, something key to address given statements in scoping reviews for national policy of insufficient evidence (Public Health England & Office for Health Improvement and Disparities, 2023). Given relatively small adjusted ORs and small PAFs of ACEs for moderate-risk or problem gambling, it is likely the gambling behaviours are a complex network of risk factors, of which ACEs are a part. Prevention strategies would need to address multiple modifiable factors alongside ACEs and trauma (Sharman et al., 2019).

These findings are in line with a body of evidence suggesting long-term potentially wide-ranging impacts of child maltreatment and between-parent violence into young adulthood, where there is already evidence for links with other adverse health behaviours and outcomes, such as smoking, substance misuse, and further violence.(Bellis, Hughes et al. 2019). Therefore, adding to the list of wider health and societal benefits of primary prevention interventions of ACEs (Asmussen et al., 2020; Gunstone & Gosschalk, 2020). Preventing ACEs through family support programmes, improved access to mental health services, and targeted social policies may have long-term benefits for reducing gambling-related harm (Mayer & Thursby, 2012). The pattern of associations, albeit imprecise, is consistent with calls for trauma-informed approaches in gambling treatment (Moxey et al., 2026), including ACE screening and tailored psychological support. Preventing ACEs through family support and mental health services may yield long-term benefits for reducing gambling harm (Moxey et al., 2026).

If these tentative associations are confirmed in future studies, they would add to arguments that gambling behaviour is shaped by broader contextual and developmental factors.. Discussions around gambling often focus on individual choice and self-control, overlooking the structural and psychological factors that contribute to gambling vulnerability. ACEs are experiences beyond an individual’s control, shaped by broader social and familial contexts, and their long-term impact on gambling risk highlights the need for systemic responses rather than individual blame. Recognising the role of childhood adversity in gambling behaviour underscores the importance of policy interventions that go beyond individual-level solutions, such as stricter gambling regulations, consumer protections, and increased investment in harm prevention and treatment strategies.

Stronger evidence of the positive associations identified here, either through combination in meta-analysis, or replication in other large longitudinal datasets, are needed. Following such work, there would be a need to better understand timing and possible mechanisms for the relationships between ACEs and gambling. There was weak evidence for associations at some time-points and not others (either only earlier or later time-points), for bullying and parental conviction. Later time-points could be explored when available in ALSPAC, to provide evidence to generate hypotheses about how particular ACEs manifest as later adverse outcomes.

### 5.4 Conclusions

The findings of our study could have important implications for both public health and gambling policy. The UK Government’s Gambling Act Review White Paper 2023 concluded that there was insufficient evidence to establish ACEs as a predictor of gambling problems (Public Health England & Office for Health Improvement and Disparities, 2023). Our study provides suggestive but imprecise longitudinal evidence of a positive association between certain ACEs and moderate risk of problem gambling, reinforcing the need to consider childhood adversity in gambling prevention and treatment efforts. Given the high prevalence of ACEs and their well-documented impact on behavioural and mental health outcomes (Bellis et al., 2019; Felitti et al., 1998), early intervention strategies that reduce exposure to childhood adversity could mitigate, amongst other outcomes, gambling-related harm. Given the uncertainty around these estimates, further research is needed to clarify whether these associations exist and, if so, through what mechanisms they might arise. Overall, although many point estimates were in the direction of increased risk, the wide and often null-spanning confidence intervals mean that these findings should be viewed as exploratory and hypothesis-generating rather than conclusive. Given these considerations, our null/weak dose–response findings should be viewed as tentative and may reflect measurement and power limitations as much as underlying aetiology.

## Data Availability

ALSPAC data access is through a system of managed open access. The <u>ALSPAC access policy (PDF, 627kB)</u> describes the process of accessing the data and samples in detail, and outlines the costs associated with doing so. You can <u>submit your research proposal</u> for consideration by the ALSPAC Executive Committee. You will receive a response within 10 working days to advise you whether your proposal has been approved. If you have any other questions about accessing data, please.

## Acknowledgements

We are extremely grateful to all the families who took part in this study, the midwives for their help in recruiting them, and the whole ALSPAC team, which includes interviewers, computer and laboratory technicians, clerical workers, research scientists, volunteers, managers, receptionists and nurses (22).

## Statement on use of Generative Artificial Intelligence (AI)

When making updates to the manuscript, the first author used Copilot version bizchat.20260316.49.2 to improve the clarity and conciseness of the manuscript’s wording. All ideas, analyses, and interpretation are the author’s own

## Footnotes

1 Problem Gambling is the official term used in the Problem Gambling Severity Index (PGSI),but there is debate about the individualising nature of this term, with a suggested move to ‘people who experience gambling problems’.

## References

Allami, Y., Hodgins, D. C., Young, M., Brunelle, N., Currie, S., Dufour, M., Flores-Pajot, M. C., & Nadeau, L. (2021). A meta-analysis of problem gambling risk factors in the general adult population. Addiction, 116(11), 2968–2977. 10.1111/add.15449

Amrhein, V., Greenland, S., & McShane, B. (2019). Scientists rise up against statistical significance. Nature, 567(7748), 305–307. 10.1038/d41586-019-00857-9

Asmussen, K., Fischer, F., Drayton, E., & McBride, T. (2020). Adverse childhood experiences: What we know, what we don’t know, and what should happen next (Summary). In Early Interv Found (pp. 128–128).

Bellis, M., Hughes, K., Ford, K., Rodriguez, G., Sethi, D., & Passmore, J. (2019). Life course health consequences and associated annual costs of adverse childhood experiences across Europe and North America : a systematic review and meta-analysis. Lancet Public Health, 4(10), 517–528. 10.1016/S2468-2667(19)30145-8

Bick, J., & Nelson, C. A. (2016). Early adverse experiences and the developing brain. Neuropsychopharmacology, 41(1), 177–196.

Blakemore, S., & Choudhury, S. (2006). Development of the adolescent brain : implications for executive function and social cognition. J Child Psychol Psychiatry, 3, 296–312.

Blaszczynski, A., Swanton Thomas, B., & Gainsbury, S. M. (2020). Avoiding use of stigmatising descriptors in gambling studies. International Gambling Studies, 20(3), 369–372. 10.1080/14459795.2020.1808774

Boyd, A., Fraser, A., Golding, J., Lawlor, D. A., Henderson, J., Ring, S., Ness, A., Molloy, L., & Davey, S. (2013). Cohort Profile: The ‘Children of the 90s’; the index offspring of The Avon Longitudinal Study of Parents and Children (ALSPAC). International Journal of Epidemiology, 42, 111–127.

Boyd, A., Golding, J., Macleod, J., Lawlor, D. A., Fraser, A., Henderson, J., Molloy, L., Ness, A., Ring, S., & Davey Smith, G. (2013). Cohort Profile: the ‘children of the 90s’--the index offspring of the Avon Longitudinal Study of Parents and Children. Int J Epidemiol, 42(1), 111–127. 10.1093/ije/dys064

Bristow, L., Afifi, T., Salmon, S., & Katz, L. (2022). Risky Gambling Behaviors: Associations with Mental Health and a History of Adverse Childhood Experiences (ACEs). J Gambl Stud, 38(3), 699–716.

Chambers, R., & Potenza, M. (2003). Neurodevelopment, Impulsivity, and Adolescent Gambling and Adolescent Gambling. J Gambl Stud, 19, 53–84.

Crick, D., Halligan, S., Howe, L., Lacey, R., Khandaker, G., Burgner, D., Herbert, A., Suderman, M., Anderson, E. L., & Fraser, A. (2022). Associations between Adverse Childhood Experiences and the novel inflammatory marker glycoprotein acetyls in two generations of the Avon Longitudinal Study of Parents and Children birth cohort. Brain Behav Immun, 100, 112–130. 2022;100:112–20. Available from: 10.1016/j.bbi.2021.11.001

Delfabbro, P., Lahn, J., & Grabosky, P. (2006). It’s Not What You Know, but How You Use It : Statistical Knowledge and Adolescent Problem Gambling. J Gambl Stud, 22(2), 179–193. 10.1007/s10899-006-9009-5

Dennie, E., Herbert, A., Sallis, H., & Rossi, R. (2023, 2023/9//). Associations of Adverse Childhood Experiences (ACEs) and gambling: a scoping review. Retrieved 11th June 2025 from https://osf.io/m4yhx/

Dowling, A., Shandley, K., Oldenhof, E., Youssef, G. J., Thomas, S. A., Frydenberg, E., & Jackson, C. The intergenerational transmission of problem gambling: The mediating role of parental psychopathology. Addictive behaviors, 59, 12–17. https://www.sciencedirect.com/science/article/abs/pii/S0306460316300958?via%3Dihub

Emond, A., & Griffiths, M. (2020). Gambling in children and adolescents. Br Med Bull, 136(1), 21–29. 10.1093/bmb/ldaa027

Emond, A., Griffiths, M. D., & Hollén, L. (2020). Problem gambling in early adulthood: A population-based study. International Journal of Mental Health and Addiction, 20(2), 754–770. 10.1007/s11469-020-00401-1

Espeleta, H., Brett, E., Ridings, L., Leavens, E., & Mullins, L. (2018). Childhood adversity and adult health-risk behaviors: Examining the roles of emotion dysregulation and urgency. Child Abus Negl, 82, 92–101.

Felitti, V. J., Anda, R. F., Nordenberg, D., Williamson, D. F., Spitz, A. M., Edwards, V., Koss, M. P., & Marks, J. S. (1998). Relationship of Childhood Abuse and Household Dysfunction to Many of the Leading Causes of Death in Adults: The Adverse Childhood Experiences (ACE) Study. American Journal of Preventive Medicine, 14(4), 245–258. 10.1016/S0749-3797(98)00017-8

Ferris J, W. H. (2001). The Canadian Problem Gambling Index : Final report. https://www.greo.ca/Modules/EvidenceCentre/files/Ferris%20et%20al(2001)The_Canadian_Problem_Gambling_Index.pdf

Ford, K., Davis, A. R., Di, L., Hardcastle, K., Hughes, K., & Edwards, S. (2019). The evidence base for routine enquiry into adverse childhood experiences : A scoping review. In Child Abuse Negl (Vol. 91, pp. 131–146).

Fraser, A., Macdonald-wallis, C., Tilling, K., Boyd, A., Golding, J., & Davey smith, G. (2013). Cohort profile: The avon longitudinal study of parents and children: ALSPAC mothers cohort. Int J Epidemiol, 42(1), 97–110.

Gambling Commission. (2024). Statistics on gambling participation – Year 1, wave 1: Official statistics. https://www.gamblingcommission.gov.uk/statistics-and-research/publication/statistics-on-gambling-participation-annual-report-year-1-2023-official.

Glasgow, G. C. N. C. f. S. R. U. o. (2024). Gambling Survey for Great Britain – Annual Report 2023. Gambling Commission. https://eprints.gla.ac.uk/330642/1/330642.pdf

Gunstone, B., & Gosschalk, K. (2020). Gambling Treatment and Support On behalf of GambleAware. Retrieved 11th June 2025 from https://assets.ctfassets.net/j16ev64qyf6l/2SXwXymIXgdAB6T30jLEuN/a33b04a4197a77af2f97c4e8b8b4cadf/gambling-treatment-and-support.pdf

Harris, P. A., Taylor, R., Thielke, R., Payne, J., Gonzalez, N., & Conde, J. G. (2009). Research electronic data capture (REDCap)--a metadata-driven methodology and workflow process for providing translational research informatics support. J Biomed Inform, 42(2), 377–381. 10.1016/j.jbi.2008.08.010

Houtepen, L., Heron, J., Suderman, M., Tilling, K., & Howe, L. (2018). Adverse childhood experiences in the children of the avon longitudinal study of parents and children (ALSPAC). Wellcome Open Res, 30(3), 106.

Howe, L., Tilling, K., Galobardes, B., & Lawlor, D. (2013). Loss to follow-up in cohort studies: bias in estimates of socioeconomic inequalities. Epidemiology, 24(1), 1–9. 10.1097/EDE.0b013e31827623b1

Hughes, K., Bellis, M. A., Hardcastle, K. A., Sethi, D., Butchart, A., Mikton, C., Jones, L., & Dunne, M. P. (2017). The effect of multiple adverse childhood experiences on health: a systematic review and meta-analysis. The Lancet Public Health, 2(8), e356–e366. 10.1016/S2468-2667(17)30118-4

Lacey, R., & Minnis, H. (2019). Practitioner Review: Twenty years of research with adverse childhood experience scores – Advantages, disadvantages and applications to practice. J Child Psychology and Psychiatry, 61(2), 116–130. 10.1111/jcpp.13135

Lane, W., Sacco, P., Downton, K., Ludeman, E., Levy, L., & Tracy, J. K. (2016). Child maltreatment and problem gambling: A systematic review. Child Abuse Negl, 58, 24–38. 10.1016/j.chiabu.2016.06.003

Lereya, S. T., Copeland, W. E., Costello, E. J., & Wolke, D. (2015). Adult mental health consequences of peer bullying and maltreatment in childhood: two cohorts in two countries. Lancet Psychiatry, 2(6), 524–531. 10.1016/s2215-0366(15)00165-0

Mayer, L. M., & Thursby, E. (2012). Adolescent Parents and Their Children: A Multifaceted Approach to Prevention of Adverse Childhood Experiences (ACE). Journal of Prevention & Intervention in the Community, 40(4), 304–312. 10.1080/10852352.2012.707448

Moxey, M., Dennie, E., Rossi, R., Sallis, H., & Herbert, A. (2026). Exploring adverse childhood experiences (ACEs) among those seeking treatment for gambling problems: A mixed-methods study. Addict Behav, 173, 108535. 10.1016/j.addbeh.2025.108535

National Institute for Health and Care Excellence. (2025, 2025/1//). Gambling-related harms: identification, assessment and management. Retrieved 11th June 2025 from https://www.nice.org.uk/guidance/NG248

NHS Digital. (2021). Health Survey for England, 2021 part 2. Retrieved 11th June 2025 from https://digital.nhs.uk/data-and-information/publications/statistical/health-survey-for-england/2021-part-2/gambling#introduction

NHS Digital. (2023). Health Survey for England 2021 – Part 2: Gambling Behaviour. NHS Digital. https://digital.nhs.uk/data-and-information/publications/statistical/health-survey-for-england/2021-part-2/gambling

Northstone, K., Lewcock, M., Groom, A., Boyd, A., Macleod, J., & Timpson, N. (2019). The Avon Longitudinal Study of Parents and Children (ALSPAC): an update on the enrolled sample of index children in 2019. Wellcome Open Res, 51–51.

Poole, J., Kim, H., Dobson, K., & Hodgins, D. (2017). Adverse Childhood Experiences and Disordered Gambling: Assessing the Mediating Role of Emotion Dysregulation. Journal of Gambling Studies, 33, 1187–1200. https://link.springer.com/article/10.1007/s10899-017-9680-8

Public Health England, & Office for Health Improvement and Disparities. (2023). Risk factors for gambling and harmful gambling: an umbrella review. A review of systematic reviews and meta-analyses. Retrieved 11th June 2025 from https://www.gov.uk/government/publications/gambling-related-harms-evidence-review/gambling-related-harms-evidence-review-summary--2

Rossi, R., & Nairn, A. (2024). Priming Young Minds: The Appeal of Gambling Advertising to Children and Young People. Journal of the Association for Consumer Research, 9(2), 187–199. 10.1086/729290

Rossi, R., & Nairn, A. (2025). Clearly (not) identifiable – The recognisability of gambling content marketing. International Journal of Market Research, 67(1), 33–52. 10.1177/14707853241292953

Rubin, D. B. (1987). Multiple imputation for nonresponse in surveys. Wiley.

Russell, A. M. T., Browne, M., Hing, N., Visintin, T., Begg, S., Rawat, V., & Rockloff, M. (2022). Stressful Life Events Precede Gambling Problems, and Continued Gambling Problems Exacerbate Stressful Life Events; A Life Course Calendar Study. J Gambl Stud, 38(4), 1405–1430. 10.1007/s10899-021-10090-7

Sharman, S., Butler, K., & Roberts, A. (2019). Psychosocial risk factors in disordered gambling: A descriptive systematic overview of vulnerable populations. Addictive behaviors, 99, 106071. 10.1016/j.addbeh.2019.106071

Smith, J., Wright, S., Dighton, G., Dymond, S., & Torrance, J. (2025). The Influence of Family on Gambling Behaviours: A Rapid Review of Emergent Literature. International Journal of Mental Health and Addiction. 10.1007/s11469-025-01505-2

Soares, S., Rocha, V., Kelly-Irving, M., Stringhini, S., & Fraga, S. (2021). Adverse Childhood Events and Health Biomarkers: A Systematic Review. Front Public Heal, 19(9), 649–825.

World Health Organisation. (2020, 2020/1//). Adverse Childhood Experiences International Questionnaire (ACE-IQ). Retrieved 11th June 2025 from https://www.who.int/publications/m/item/adverse-childhood-experiences-international-questionnaire-%28ace-iq%29

