## Supplementary Files for "Longitudinal associations between adverse childhood experiences with moderate-risk to problem gambling in young adulthood: A prospective UK cohort study"

**Supplementary materials**

**Box S1. Formula for estimating Population Attributable Fractions**

We used the following formula:

$$\hat{PAF}=\frac{\hat{P}(A=1|Y=1)(e^{\hat{\beta}_{1}}-1)}{e^{\hat{\beta}_{1}}}\times100$$

where $\hat{P}(A=1|Y=1)$ is the estimated prevalence of the ACE in question among those with a positive outcome (moderate-risk+ gambling) and $e^{\hat{\beta}_{1}}$ is the estimated aOR of moderate-risk+ gambling at ages 17, 20, and 24 years for each of the 10 ACEs (Ferguson et al., 2019). PAFs provide an alternative interpretation of aORs in the context of this study’s ACE prevalence, as they are a function of both aOR and prevalence. In other words, PAFs indicate the proportion of gambling cases in this population that could be attributed to each ACE—suggesting the potential number of cases that might be prevented if that ACE were eliminated.

**Table S1. Crude estimates for the relationship between Adverse Childhood Experiences and moderate-risk+ gambling at ages 17, 20, and 24^a^**

|  | **Age 17** |  |  | **Age 20** |  |  | **Age 24** |  |  |
| --- | --- | --- | --- | --- | --- | --- | --- | --- | --- |
| **Adverse Childhood Experience** | **OR** | **(CI)** | **P-Value** | **OR** | **(CI)** | **P-Value** | **OR** | **(CI)** | **P-Value** |
| Physical abuse | 1.59 | (0.85 to 2.96) | 0.14 | 1.34 | (0.87 to 2.07) | 0.19 | 1.32 | (0.78 to 2.25) | 0.31 |
| Sexual abuse | 1.89 | (0.60 to 5.98) | 0.28 | 1.01 | (0.47 to 2.18) | 0.97 | 2.03 | (1.03 to 4.02) | 0.04 |
| Emotional abuse | 1.10 | (0.53 to 2.27) | 0.8 | 1.23 | (0.81 to 1.89) | 0.34 | 1.31 | (0.79 to 2.18) | 0.29 |
| Emotional neglect | 1.36 | (0.70 to 2.67) | 0.37 | 1.53 | (1.02 to 2.30) | 0.04 | 1.49 | (0.91 to 2.44) | 0.12 |
| Bullying | 1.54 | (0.89 to 2.68) | 0.12 | 1.41 | (0.97 to 2.06) | 0.07 | 1.25 | (0.79 to 1.98) | 0.34 |
| Violence between parents | 1.31 | (0.63 to 2.73) | 0.48 | 1.28 | (0.80 to 2.06) | 0.31 | 1.29 | (0.73 to 2.27) | 0.38 |
| Household substance abuse | 0.91 | (0.35 to 2.36) | 0.84 | 0.81 | (0.43 to 1.52) | 0.51 | 0.91 | (0.43 to 1.89) | 0.8 |
| Parental mental health | 1.02 | (0.61 to 1.71) | 0.94 | 1.10 | (0.78 to 1.57) | 0.58 | 0.95 | (0.62 to 1.47) | 0.83 |
| Parent convicted of an offence | 2.00 | (0.89 to 4.49) | 0.09 | 1.29 | (0.67 to 2.46) | 0.45 | 1.17 | (0.53 to 2.54) | 0.7 |
| Parental separation | 1.2 | (0.65 to 2.23) | 0.56 | 0.79 | (0.49 to 1.27) | 0.34 | 1.27 | (0.79 to 2.02) | 0.32 |
| ACE score (vs. 0) |  |  |  |  |  |  |  |  |  |
| 1 | 0.96 | (0.44 to 2.12) | 0.92 | 0.82 | (0.47 to 1.42) | 0.47 | 1.24 | (0.69 to 2.21) | 0.47 |
| 2-3 | 1.53 | (0.51 to 4.57) | 0.45 | 0.89 | (0.52 to 1.54) | 0.68 | 1.05 | (0.55 to 2.02) | 0.87 |
| 4+ | 2.11 | (0.55 to 8.07) | 0.28 | 0.91 | (0.41 to 2.00) | 0.81 | 1.25 | (0.50 to 3.09) | 0.63 |

a. This table displays the results of 11 separate logistic regression models, pooled across 20 imputed datasets.

**Table S2. Adjusted estimates for the relationship between Adverse Childhood Experiences and moderate-risk+ gambling at ages 17, 20, and 24^a^**

|  | **Age 17** |  |  | **Age 20** |  |  | **Age 24** |  |  |
| --- | --- | --- | --- | --- | --- | --- | --- | --- | --- |
| **Adverse Childhood Experience** | **OR** | **(CI)** | **P-Value** | **OR** | **(CI)** | **P-Value** | **OR** | **(CI)** | **P-Value** |
| Physical abuse | 1.21 | (0.57 to 2.57) | 0.62 | 1.36 | (0.84 to 2.20) | 0.21 | 1.26 | (0.70 to 2.28) | 0.44 |
| Sexual abuse | 2.29 | (0.5 to 10.47) | 0.28 | 2.26 | (0.93 to 5.48) | 0.07 | 4.19 | (1.79 to 9.78) | <0.01 |
| Emotional abuse | 0.99 | (0.44 to 2.22) | 0.98 | 1.29 | (0.81 to 2.05) | 0.28 | 1.53 | (0.87 to 2.68) | 0.14 |
| Emotional neglect | 1.12 | (0.49 to 2.54) | 0.79 | 1.73 | (1.09 to 2.75) | 0.02 | 1.58 | (0.89 to 2.81) | 0.12 |
| Bullying | 1.61 | (0.86 to 3.01) | 0.14 | 1.32 | (0.86 to 2.01) | 0.20 | 1.04 | (0.62 to 1.75) | 0.89 |
| Violence between parents | 1.20 | (0.53 to 2.74) | 0.66 | 1.46 | (0.86 to 2.49) | 0.17 | 1.28 | (0.69 to 2.35) | 0.43 |
| Household substance abuse | 0.61 | (0.16 to 2.42) | 0.49 | 0.95 | (0.46 to 1.94) | 0.88 | 0.86 | (0.34 to 2.22) | 0.76 |
| Parental mental health | 1.23 | (0.67 to 2.25) | 0.50 | 1.20 | (0.80 to 1.79) | 0.39 | 0.89 | (0.55 to 1.45) | 0.63 |
| Parent convicted of an offence | 2.35 | (0.91 to 6.04) | 0.08 | 1.26 | (0.58 to 2.74) | 0.57 | 0.93 | (0.33 to 2.6) | 0.89 |
| Parental separation | 0.96 | (0.44 to 2.12) | 0.92 | 0.82 | (0.47 to 1.42) | 0.47 | 1.24 | (0.69 to 2.21) | 0.47 |
| ACE score (vs. 0) |  |  |  |  |  |  |  |  |  |
| 1 | 0.92 | (0.31 to 2.73) | 0.88 | 0.74 | (0.41 to 1.35) | 0.33 | 0.75 | (0.34 to 1.65) | 0.48 |
| 2-3 | 1.47 | (0.62 to 3.51) | 0.38 | 1.27 | (0.74 to 2.19) | 0.38 | 1.46 | (0.74 to 2.9) | 0.27 |
| 4+ | 1.45 | (0.49 to 4.32) | 0.50 | 1.42 | (0.73 to 2.78) | 0.30 | 1.27 | (0.54 to 2.96) | 0.58 |

a. This table displays the results of 11 separate ordinal logistic regression models, pooled across 20 imputed datasets. Each model was adjusted for the following variables: sex, maternal home ownership status, parity, maternal smoking in pregnancy, mother and father’s highest educational qualification, household social class, child ethnic background, mother at age of delivery, and mother and father’s depression scores. Evidence of association is based on 95% CIs.

**Table S3. Crude estimates for the relationship between Adverse Childhood Experiences and at-risk+ gambling at ages 17, 20, and 24^a^**

|  | **Age 17** |  |  | **Age 20** |  |  | **Age 24** |  |  |
| --- | --- | --- | --- | --- | --- | --- | --- | --- | --- |
| **Adverse Childhood Experience** | **OR** | **(CI)** | **P-Value** | **OR** | **(CI)** | **P-Value** | **OR** | **(CI)** | **P-Value** |
| Physical abuse | 1.08 | (0.75 to 1.57) | 0.68 | 1.36 | (1.05 to 1.76) | 0.02 | 1.22 | (0.91 to 1.65) | 0.19 |
| Sexual abuse | 1.09 | (0.49 to 2.39) | 0.84 | 1.23 | (0.80 to 1.89) | 0.35 | 1.36 | (0.84 to 2.20) | 0.22 |
| Emotional abuse | 0.90 | (0.62 to 1.31) | 0.60 | 1.07 | (0.84 to 1.35) | 0.60 | 1.00 | (0.72 to 1.38) | 0.99 |
| Emotional neglect | 1.45 | (1.00 to 2.10) | 0.05 | 1.25 | (0.97 to 1.61) | 0.08 | 1.20 | (0.87 to 1.65) | 0.26 |
| Bullying | 1.35 | (0.99 to 1.84) | 0.05 | 1.32 | (1.05 to 1.66) | 0.02 | 0.93 | (0.69 to 1.24) | 0.60 |
| Violence between parents | 1.10 | (0.72 to 1.67) | 0.66 | 0.99 | (0.76 to 1.28) | 0.92 | 1.40 | (1.03 to 1.90) | 0.03 |
| Household substance abuse | 1.11 | (0.67 to 1.84) | 0.69 | 0.83 | (0.59 to 1.16) | 0.27 | 1.13 | (0.78 to 1.63) | 0.52 |
| Parental mental health | 1.10 | (0.83 to 1.46) | 0.50 | 1.02 | (0.84 to 1.24) | 0.86 | 0.87 | (0.69 to 1.12) | 0.28 |
| Parent convicted of an offence | 1.21 | (0.69 to 2.15) | 0.51 | 1.28 | (0.89 to 1.84) | 0.18 | 1.00 | (0.62 to 1.61) | 0.99 |
| Parental separation | 1.08 | (0.76 to 1.53) | 0.68 | 1.09 | (0.86 to 1.39) | 0.46 | 1.35 | (1.02 to 1.78) | 0.04 |
| ACE score (vs. 0) |  |  |  |  |  |  |  |  |  |
| 1 | 1.18 | (0.75 to 1.85) | 0.47 | 1.13 | (0.85 to 1.51) | 0.41 | 0.87 | (0.60 to 1.25) | 0.45 |
| 2-3 | 1.29 | (0.85 to 1.94) | 0.23 | 1.25 | (0.94 to 1.65) | 0.13 | 1.07 | (0.76 to 1.49) | 0.70 |
| 4+ | 1.41 | (0.86 to 2.30) | 0.17 | 1.38 | (0.98 to 1.94) | 0.06 | 1.20 | (0.81 to 1.77) | 0.36 |

a. This table displays the results of 11 separate ordinal logistic regression models, pooled across 20 imputed datasets.

**Table S4. Adjusted estimates for the relationship between Adverse Childhood Experiences and at-risk+ gambling at ages 17, 20, and 24^a^**

|  | **Age 17** | | | **Age 20** | | | **Age 24** | | |
| --- | --- | --- | --- | --- | --- | --- | --- | --- | --- |
| **Adverse Childhood Experience** | **OR** | **(CI)** | **P-Value** | **OR** | **(CI)** | **P-Value** | **OR** | **(CI)** | **P-Value** |
| Physical abuse | 0.96 | (0.62 to 1.49) | 0.85 | 1.28 | (0.97 to 1.7) | 0.08 | 1.10 | (0.78 to 1.56) | 0.59 |
| Sexual abuse | 1.75 | (0.72 to 4.23) | 0.22 | 1.78 | (1.04 to 3.04) | 0.03 | 2.08 | (1.13 to 3.82) | 0.02 |
| Emotional abuse | 0.92 | (0.61 to 1.41) | 0.71 | 1.03 | (0.79 to 1.36) | 0.81 | 1.09 | (0.77 to 1.54) | 0.62 |
| Emotional neglect | 1.52 | (0.97 to 2.37) | 0.07 | 1.32 | (1.00 to 1.74) | 0.05 | 1.17 | (0.81 to 1.69) | 0.41 |
| Bullying | 1.26 | (0.89 to 1.80) | 0.20 | 1.40 | (1.08 to 1.83) | 0.01 | 0.83 | (0.6 to 1.15) | 0.27 |
| Violence between parents | 1.06 | (0.68 to 1.66) | 0.79 | 0.97 | (0.71 to 1.31) | 0.83 | 1.39 | (0.98 to 1.98) | 0.07 |
| Household substance abuse | 1.05 | (0.58 to 1.93) | 0.86 | 0.79 | (0.53 to 1.17) | 0.24 | 1.06 | (0.66 to 1.70) | 0.81 |
| Parental mental health | 1.15 | (0.83 to 1.59) | 0.4 | 1.05 | (0.84 to 1.32) | 0.66 | 0.76 | (0.57 to 1.00) | 0.05 |
| Parent convicted of an offence | 1.32 | (0.73 to 2.40) | 0.35 | 1.32 | (0.88 to 1.98) | 0.17 | 0.78 | (0.42 to 1.43) | 0.43 |
| Parental separation | 1.05 | (0.68 to 1.61) | 0.84 | 1.10 | (0.83 to 1.45) | 0.50 | 1.38 | (0.99 to 1.93) | 0.06 |
| ACE score (vs. 0) |  |  |  |  |  |  |  |  |  |
| 1 | 1.13 | (0.69 to 1.87) | 0.62 | 1.18 | (0.86 to 1.62) | 0.31 | 0.81 | (0.54 to 1.22) | 0.31 |
| 2-3 | 1.27 | (0.78 to 2.07) | 0.33 | 1.34 | (0.99 to 1.82) | 0.06 | 1.01 | (0.69 to 1.48) | 0.94 |
| 4+ | 1.39 | (0.8 to 2.44) | 0.25 | 1.44 | (0.97 to 2.12) | 0.07 | 1.09 | (0.68 to 1.73) | 0.72 |

a. This table displays the results of 11 separate ordinal logistic regression models, pooled across 20 imputed datasets. Each model was adjusted for the following variables: sex, maternal home ownership status, parity, maternal smoking in pregnancy, mother and father’s highest educational qualification, household social class, child ethnic background, mother at age of delivery, and mother and father’s depression scores. Evidence of association is based on 95% CIs.

**Table S5. Population Attributable Fractions (PAFs) for individual Adverse Childhood Experiences and at-risk+ gambling at age 17, 20 and 24 years**

| **ACE** | **Age 17** |  | **Age 20** |  | **Age 24** |  |
| --- | --- | --- | --- | --- | --- | --- |
| Physical abuse | -0.8 | (-12.3 to 6.5) | 4.7 | (-0.7 to 8.8) | 2.0 | (-6.2 to 7.8) |
| Sexual abuse | 1.9 | (-1.7 to 3.3) | 2.4 | (0.2 to 3.7) | 3.2 | (0.7 to 4.6) |
| Emotional abuse | -1.5 | (-11.9 to 5.3) | 0.6 | (-5.3 to 5.1) | 1.5 | (-5.3 to 6.3) |
| Emotional neglect | 7.1 | (-0.6 to 12.1) | 5.2 | (0 to 9.1) | 2.7 | (-4.4 to 7.6) |
| Bullying | 5.8 | (-3.6 to 12.3) | 7.9 | (2 to 12.4) | -4.4 | (-14.4 to 2.9) |
| Violence between parents | 1.1 | (-9.1 to 7.6) | -0.6 | (-7.8 to 4.6) | 6.7 | (-0.5 to 11.7) |
| Household substance abuse | 0.6 | (-8.2 to 5.4) | -2.6 | (-8.4 to 1.4) | 0.7 | (-6.1 to 4.9) |
| Parental mental health problems or suicide | 5.9 | (-9.1 to 16.7) | 2.1 | (-8.2 to 10.3) | -12.6 | (-29.4 to 0.1) |
| Parent convicted of an offence | 2.2 | (-3.3 to 5.3) | 2.1 | (-1.1 to 4.2) | -2.1 | (-10.2 to 2.3) |
| Parental separation | 1.3 | (-13.7 to 11) | 2.6 | (-5.8 to 9) | 8.7 | (-0.3 to 15.2) |
